# COVID-19 vaccine breakthrough infections among fully vaccinated Health Care Workers in Lagos, Nigeria

**DOI:** 10.1101/2022.06.22.22276765

**Authors:** David A. Oladele, Abideen Salako, James Ayorinde, Chika Onwuamah, Olagoke Usman, Rufai Abubakar, Gideon Liboro, Oluwatosin Odubela, Sunday Mogaji, Fehintola Ige, Gregory Ohihoin, Oliver Ezechi, Rosemary Audu, Richard A. Adegbola, Adedamola Dada, Tunde Salako

## Abstract

**Background:** Access to vaccines has contributed to the control of COVID-19. However, evaluation of the effectiveness of the vaccines in a setting where the vaccines were not originally tested is critically important. This study evaluates the clinical and laboratory characteristics of COVID-19 vaccine breakthrough infections among healthcare workers (HCWs).

**Methods:** A multicentre prospective study among HCWs who had two doses of the Oxford/AstraZeneca ChAdOx1-S [recombinant] (AZD1222) vaccine were followed up 24 weeks. Nasopharyngeal and oropharyngeal specimens were tested using RT-PCR for SARS-CoV-2 and positive samples were subjected to whole genome sequencing for variant assignment.

**Result:** A total of 369 HCWs were enrolled; of which 24 (6.5%) had breakthrough infections. There was equal sex distribution among the breakthrough cases. The majority were aged between 30 to 39years (37.5%), and had mild symptoms of cough, fever, headache, and nausea/vomiting (58%), with no hospitalization. Among the 24 breakthrough cases whose whole genomes were successfully sequenced, three were confirmed to be Delta B.1.617.2 variant during the 3^rd^ wave and an additional three were confirmed as omicron B.1.1.529 variant during the 4^th^ wave.

**Conclusion:** We reported vaccine breakthrough cases among fully vaccinated HCWs with the majority presenting with mild illness. Both delta and omicron variants were identified during the different epidemiologic spectrums of SARS-CoV-2. Therefore, there is a need to scale up vaccination for all front-line health workers and high-risk populations in developing countries.

## Introduction

Ever since the outbreak of Coronavirus disease (COVID-19) caused by Severe Acute Respiratory Syndrome Coronavirus 2 (SARS-COV-2), all effort has been geared toward finding an effective vaccine to curtail the spread of the virus [1]. As of 1^st^ April 2022, there were 153 candidate vaccines against SARS-CoV-2 in the clinical development phase and 196 candidate vaccines in the preclinical development phase, and a total of 8 have been granted Emergency Use Listing (EUL) by the WHO globally [2,3]. Four randomized controlled trials carried out in the United Kingdom, South Africa, and Brazil reported the safety and efficacy of the AZD1222 vaccine against SARS-CoV-2 [4]. The efficacy of AZD1222 was 63.09% against symptomatic infection [5]. However, longer dosage intervals between doses (8-12weeks) are associated with greater vaccine efficacy [4].

In the mid of the first quarter of 2021, AZD1222 arrived in Nigeria. However, due to limited doses available, it was recommended that priority be given to health care workers (HCWs) at high risk of exposure and the elderly who were considered a high-risk population for morbidity and mortality from the COVID-19 [5]. The HCWs were also considered a high-risk group because an asymptomatic infected person may cause a horizontal spread of the virus during the cause of work and the vaccine has been shown to reduce the incidence of asymptomatic infection and the associated infectivity [6].

However, breakthrough infections have been reported globally among the HCWs that have been fully vaccinated [7,8]. A breakthrough infection was defined as the detection of SARS-CoV-2 on RT-PCR assay performed 11 or more days after receipt of the second dose of ChAdOx1-S if no explicit exposure or symptoms had been reported during the first 6 days. Some studies have shown that breakthrough cases of COVID-19 are associated with asymptomatic infection or milder symptoms. Other studies have implicated a different variant of SARS-COV-2 following vaccination (Alpha/UK variant B.1.1.7, Beta/South African B.1.351, Gamma/Brazilian variant, and the Kappa/Delta B.1.617.1/B.1.617.2 variants) as the cause of the breakthrough infection [9]. Therefore, vaccinated individuals are still advised to maintain and strengthen public health measures (masking, physical distancing, hand washing, respiratory and cough hygiene, avoiding crowds, and ensuring good ventilation), especially when in confined spaces to adequately protect themselves and loved ones [10]. There is a paucity of data on COVID-19 vaccine breakthrough cases in low and middle-income countries.

Following the rollout of the Oxford Astrazeneca ChAdOx1-S [recombinant] vaccine in Lagos State, Nigeria, we undertook a study to evaluate SARS-CoV-2 breakthrough infections among fully vaccinated HCWs in two healthcare centers in Lagos and report our findings as part of a study designed to study the effectiveness of COVID-19 vaccination in HCWs.

## Methods

### Study locations and enrolment procedure

The study was conducted among the fully vaccinated HCWs of two health facilities in Lagos: The Nigerian Institute of Medical Research, (NIMR) Yaba, and the Federal Medical Center, (FMC) Ebute Metta). A total of 50 and 319 HCWs at FMC and NIMR respectively who had received two doses of AZD1222 at intervals of 12 weeks were followed up 24 weeks after receiving the second dose (Fig 1: Study timeline).

**Figure.**
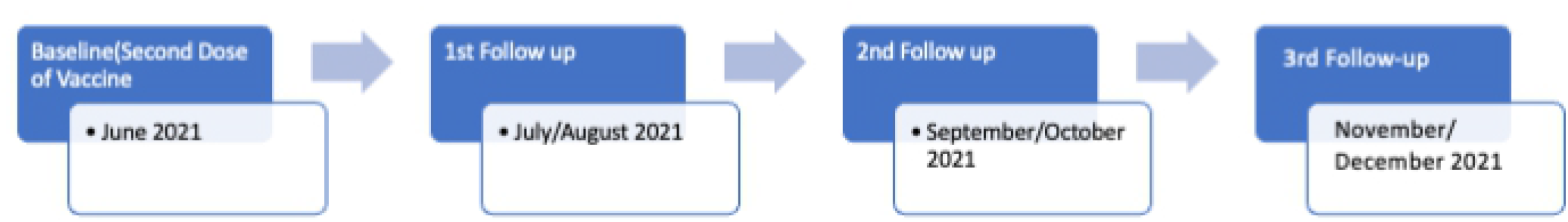

Healthcare workers who consented to be part of the COVID-19 vaccine effectiveness study and who received 2 doses of the COVID-19 vaccines at the 2 facilities were recruited into the study. Participants with confirmed breakthrough infection were interviewed using an adapted questionnaire for public health investigation of COVID-19 vaccine breakthrough cases [11].

Nasopharyngeal and oropharyngeal swabs of all vaccinated HCWs were taken at 4 to 6 weeks, 8 to 12 weeks, and 16-24 weeks following the second dose of the vaccine and tested for the presence of SARS-CoV-2 using reverse-transcriptase– polymerase-chain-reaction (RT-PCR) assay. HCWs who had COVID-19-related symptoms or were exposed to infected persons voluntarily presented to the NIMR for testing. We identified a breakthrough infection as the detection of SARS-CoV-2 infection in individuals, who have been fully vaccinated against COVID-19. The identified breakthrough cases were interviewed using a questionnaire for investigation of COVID-19 vaccine breakthrough cases adapted from the Centers for Disease Control and Prevention (CDC) protocol, we were able to describe the clinical profile of those with breakthrough infections.

### Laboratory Methods

Swab samples collected were tested using Cobas® SARS-CoV-2 qualitative assay on Cobas® 6800 System (Roche Molecular Systems, Rotkreuz, Switzerland) for the detection of the 2019 novel coronavirus (SARS-CoV-2) RNA [12].

Briefly, oropharyngeal and nasopharyngeal swabs were collected by trained personnel, and RT-PCR testing was performed with the use of the **Cobas®** SARS-CoV-2. **Cobas®** SARS-CoV-2 testing is based on fully automated sample preparation (nucleic acid extraction and purification) followed by PCR amplification and detection. Automated data management was performed by the **Cobas®** 6800/8800 software which assigns test results for all tests.

Selective amplification of target nucleic acid from the sample is achieved by the use of target-specific forward and reverse primers for ORF1 a/b non-structural region unique to SARS-CoV-2. Additionally, a conserved region in the structural protein envelope E-gene was chosen for pan-Sarbecovirus detection. The pan-Sarbecovirus detection sets also detected the SARS-CoV-2 virus. Selective amplification of RNA Internal Control is achieved by the use of non-competitive sequence-specific forward and reverse primers that have no homology with the coronavirus genome.

Complete genome sequencing of SARS-CoV-2 positive samples was done using the ATOPlex RNA Universal Library Preparation module developed by MGI Co. Ltd (China) [13]. In brief, extracted viral RNA samples were converted to cDNA which was used to generate tiling PCR amplicons. Dual barcodes for each specimen were enzymatically tagged to end-repaired amplicons. Cleaned libraries were pooled, denatured, circularized, and amplified using rolling circle amplification (RCA) to generate DNA nanoballs (DNB). Normalized DNB libraries were sequenced using paired end DNBSEQ-G50RS High-throughput Sequencing kit with a large flow cell (FCL PE100) [14].

### Ethical consideration

Ethical approval for the study was obtained from the Institutional Review Board of the Nigerian Institute of Medical Research (IRB-21-040) and written informed consent was obtained from all the study participants prior to enrolment in the study.

### Data processing and Analysis

The MGI SARS-CoV-2 bioinformatics analysis pipeline in GitHub [15] was used to perform the quality assessment of the de-multiplexed reads and to generate consensus sequences in FASTA format. For all analyses, default parameters were used. The complete genome sequences of each of the SARS-CoV-2 isolates were submitted to PANGOLIN (web-based) for lineage assignment [16].

Data management and analysis were done using SPSS version 23. Descriptive statistic was done to adequately document the clinical and laboratory characteristics of the breakthrough cases.

## Results

The socio-demographic characteristics of persons with breakthrough infections are shown in table 1. Among 369 fully vaccinated HCWs in the two health facilities, 24 (6.5%) breakthrough cases were detected with equal sex distribution (50%) and the age ranging from 30 to 39 years with a mean of 38.7 years.

**Table 1:** Socio-demographic characteristics of the 24 persons with breakthrough infection

| Variable | Number (%) |
| --- | --- |
| Age (years) |  |
| 20-29 | 5 (20.8) |
| 30-39 | 9 ( 37.5) |
| 40-49 | 7 (29.2) |
| 50-65 | 3 (12.5) |
| Mean Age | 38.7 |
| Sex |  |
| Female | 12 (50.0) |
| Male | 12 (50.0) |
| Place of practice |  |
| NIMR | 18 (75.0) |
| FMC | 6 (25.0) |
| Job description of the HCW |  |
| Laboratory staff | 8 (33.3) |
| Admin staff | 4 (16.7) |
| Data officer | 3 (12.5) |
| COVID-19 test centre staff | 2 (8.3) |
| Works/maintenance | 2 (8.3) |
| Doctor | 1 (4.2) |
| Pharmacist | 1 (4.2) |
| Psychologist | 1 (4.2) |
| Physiotherapist | 1 (4.2) |
| Biotech(MBT) staff | 1 (4.2) |

A review of the job description of participants shows that 8 of the breakthrough cases work at the various laboratory complexes, 4 hold various administrative duties, 3 were data officers managing hospital databases, 2 were COVID-19 drive-through test center staff, and 2 were staff of maintenance department. Others did include a medical doctor, a pharmacist, a physiotherapist, and a psychologist.

Evaluation of the spectrum of COVID-19 symptoms among the 24 breakthrough cases, revealed that 14 had mild symptoms and 10 participants were asymptomatic (Table 2). Some of the reported symptoms include cough (41.7%), fever (33.3%), headache (29.2%), chills (12.5%) nausea and vomiting (12.5%), and fatigue (8.3%). Most of the reported symptoms did not require hospital admission though about 16.7% of the symptomatic cases required outpatient care. Others were treated at home using over-the-counter medications. There was no history of hospitalization, need for breathing support, or oxygen use, and no history of death among the breakthrough cases. Also, none of the symptomatic cases had a history of immunosuppressive disease or are on immunosuppressive therapy.

**Table 2:** Clinical symptoms of the participants with breakthrough infection

| Clinical symptoms | Frequency(percentage) |
| --- | --- |
| Fever | 8 (33.3) |
| Chills | 3 (12.5) |
| Rigor | 0 (0.0) |
| Myalgia | 2 (8.3) |
| Headache | 7 (29.2) |
| Sore throat | 2 (8.3) |
| Nausea and vomiting | 3 (12.5) |
| Diarrhea | 1 (4.2) |
| Fatigue | 2 (8.3) |
| Congestion/ runny nose | 2 (8.3) |
| Cough | 10 (41.7) |
| Chest pain | 1 (4.2) |
| New taste disorder | 1 (4.2) |

On follow-up assessment, all the breakthrough cases recovered fully the following self-isolation for 2 weeks and a repeat of the nasopharyngeal and oropharyngeal swab for RT PCR for each one was all negative. Nine of the samples from the breakthrough cases reported during the second follow-up period (3^rd^ wave of COVID-19 infection in Nigeria) were sequenced at the Central Laboratories at NIMR using the next-generation sequencing. Lineage assignment using Pangolin did show that three of the breakthrough cases are B.1.617.2 Delta variant. Also, six of the breakthrough cases reported during the 3^rd^ follow-up period (Corresponding to the 4^th^ wave of COVID-19 in Nigeria) were sequenced and three were confirmed as Omicron B.1.1.529 variant while the remaining three samples could not be sequenced because the CT values were above 34.

## Discussion

Since the emergency use listing (EUL) of different COVID-19 vaccines by the WHO, there has been a considerable drop in the number of persons with severe disease, hospitalization, and death [17]. Several studies have reported a reduction in the neutralization potential of the sera of individuals vaccinated, hence the risk of breakthrough infection [18]. This study, therefore, describes the clinical and laboratory spectrum of COVID-19 vaccine breakthrough cases in Lagos, Nigeria. There were 24 vaccine breakthrough infections among the 369 fully vaccinated health care workers enrolled during the period of this study. Equal sex distribution was observed (50.0%) among participants with breakthrough infection and the cases occurred among participants who work as a laboratory staff (33.3%), administrative staff (16.7%), data officers (12.5%), COVID-19 test center staff (8.3%) and maintenance department staffs (8.3%).

The findings in our study are different from the reported predominant health workers with vaccine breakthrough cases in previous works [19-24]. For instance, a study from Vietnam reported a greater proportion (8%) of vaccine breakthrough cases among healthcare workers [25].

The majority of the HCWs had mild symptoms which do not require hospitalization, ranging from cough (41.7%), fever (33.3%), and headache (29.2%). This finding is similar to another report of COVID-19 vaccine breakthrough cases reported from China in which most of the cases were either asymptomatic or with mild symptoms not requiring hospitalization [25]. A similar study in Israel showed a percentage breakthrough case of 2.6% (out of 1,497 HCWs tested) among HCWs, the majority having milder symptoms. However, a few were noted to have required hospitalization (9.8%) with full recovery after 6 weeks [23]. Also, studies in Kerala and Delhi, India reported mild symptoms among the COVID-19 vaccine breakthrough cases in their setting. [24]. Thus, the vaccine reduces the severity of the infection, the need for hospitalization, and mortality. Therefore, enhancing the vaccination drive and immunizing the populations quickly would be an important strategy to prevent further morbidity and mortality from the COVID-19 and would reduce the burden on the health care system.

The Delta variant has been designated as a variant of concern due to increased transmission and higher immune evasion [19]. The emergence of a variant of concerns has led to an upsurge in COVID-19 cases and the second wave of pandemics globally. Incidentally, several countries have reported COVID-19 breakthrough infections even after completion of the full vaccination schedule [20,21]. Over 10,000 breakthrough infections after completion of the full course of vaccination, representing less than 1 % of the vaccinated population have been reported in the USA [22]. Overall, breakthrough infections were seen in a smaller percentage of the total vaccinated population. Our study report complete-genome sequences among the vaccine breakthrough cases seen during the 3^rd^ wave and 4^th^ wave of the COVID-19 infection in Nigeria in which three cases were Delta variant (B.1.617.2) during the 3^rd^ wave and 3 were omicron during the 4^th^ wave. This is similar to reports from other parts of the world in which the delta and the omicron variants were reported as the predominant variants in the 3rd and 4^th^ wave of COVID-19 respectively [26-28].

The strength of the study includes the fact that it was carried out among high-risk individuals(HCWs) and the availability of PCR testing and genomic sequencing facility which helped in the diagnosis and characterization of the prevalent SARS-CoV-2 variants in Nigeria. To the best of our knowledge, this is the first report of vaccine breakthrough infections among HCWs after completion of the full course of COVID-19 vaccination in Nigeria. Our study has several limitations including lacking the capability to carry out real-time neutralizing antibody tests among the vaccine breakthrough cases and the inability to perform complete genome sequencing for all the vaccine breakthrough cases.

## Conclusions

With the availability of the COVID-19 vaccine in Nigeria, the front-line health worker was the priority group for vaccination. We reported vaccine breakthrough cases among fully vaccinated HCWs, and the spectrum of the clinical presentation was asymptomatic to mild cases without any hospitalization and mortality. There is a need to scale up vaccination of all front-line health workers in developing countries as well as other high-risk populations.

## Data Availability

The datasets generated during and/or analysed during the current study are available from the corresponding author on reasonable request through the IRB of the Nigerian Institute of Medical Research

## Authors contribution

Designing research studies: DAO,RA,RAA

Laboratory analysis: JA,CO,GL,FI

Acquiring data: DAO,OU,RA,GL,OO,SM

Analysing data: DAO,GO,RA,RA

Writing the manuscript: DAO,JA,OE,RA,RAA,AD,BS

